# High-Plex and High-throughput Digital Spatial Profiling of non-small-cell lung cancer (NSCLC)

**DOI:** 10.1101/2020.07.22.20160325

**Authors:** James Monkman, Touraj Taheri, Majid Ebrahimi Warkiani, Connor O’leary, Rahul Ladwa, Derek Richard, Ken O’ Byrne, Arutha Kulasinghe

## Abstract

Profiling the tumour microenvironment(TME) has been informative in understanding the underlying tumour-immune interactions. Multiplex immunohistochemistry(mIHC) coupled with molecular barcoding technologies have revealed greater insights into the TME. In this study, we utilised the Nanostring GeoMX™ Digital Spatial Profiler (DSP) platform to profile a NSCLC tissue microarray for protein markers across immune cell profiling, immuno-oncology(IO) drug target, immune activation status, immune cell typing, and pan-tumour protein modules. Regions of interest(ROIs) were selected that described tumour, TME and normal adjacent tissue(NAT) compartments. Our data revealed that paired analysis (n=18) of patient matched compartments indicated that the TME was significantly enriched in CD27, CD3, CD4, CD44, CD45, CD45RO, CD68, CD163, and VISTA relative to tumour. Unmatched analysis indicated that the NAT(n=19) was significantly enriched in CD34, fibronectin, IDO1, LAG3, ARG1 and PTEN when compared to the TME(n=32). Univariate Cox proportional hazards indicated that the presence of cells expressing CD3(HR:0.5, p=0.018), CD34(HR:0.53, p=0.004) and ICOS (HR:0.6, p=0.047) in tumour compartments were significantly associated with improved overall survival(OS). We implemented both high-plex and high-throughput methodologies to the discovery of protein biomarkers and molecular phenotypes within biopsy samples and demonstrate the power of such tools for a new generation of pathology research.

**Conflict of interest statement:** The authors have declared that no conflict of interest exists.

## Introduction

Non-small-cell lung cancer (NSCLC) accounts for 85% of lung cancers and is the leading cause of cancer related deaths ^1^. Patients are often diagnosed at an advanced stage, where the immediate prognosis is poor, resulting in a 5-year survival rate of less than 20% ^2,3^. With the emerging success of immune checkpoint blockade leading to durable responses and prolonged survival in 15-40% of cases, there is now a need for predictive biomarkers to guide patient selection for targeted therapies ^4^. The use of comprehensive tumoural information to inform clinical decision making is becoming increasingly important ^5-9^. Studies in the tumour microenvironment (TME) have revealed that a high degree of T-cell infiltration into the tumour provides fertile grounds for effective immunotherapies ^10^. As such, the immune contexture (type, density, location, phenotypic and functional profile of immune cells) has been used to understand a greater depth of the tumour-immune cell interactions which may provide cues into predictive biomarkers of response to immune checkpoint therapy (anti PD-1/PD-L1) ^11,12^.

Whilst traditional immunohistochemistry (IHC) techniques allow for spatial profiling of cells in the tumour, this is often lost when tumours are analysed using bulk tissue genomic approaches. Moreover, the actual cellular proportions, cellular heterogeneity and deeper spatial distribution are lacking in characterisation. Spatial and immunological composition with cellular status can aid in identifying micro-niches within the TME ^13^. The classification of the immune context within the TME lays the foundation to addressing how the immunological composition and status (activated/suppressed) may dictate response to therapy. Therefore, to address this need, imaging and tissue sampling is required simultaneously to analyse tumour tissue and immune proteins with spatial resolution.

In this study we used the Nanostring GeoMX™ Digital Spatial Profiler (DSP) to measure compartment specific expression of proteins across immune cell profiling, immuno-oncology (IO) drug target, immune activation status, immune cell typing, and pan-tumour protein modules. We found that in paired analysis of matched compartments, the TME was enriched for CD27, CD3, CD4, CD44, CD45, CD45RO, CD68, CD163 and VISTA relative to the tumour regions. Unmatched analysis indicated that the NAT (n=19) was significantly enriched in CD34, fibronectin, IDO1, LAG3, ARG1 and PTEN when compared to TME. Univariate Cox proportional hazard analysis indicated that the presence of cells expressing CD3 (HR:0.5, p=0.018), CD34 (HR:0.53, p=0.004) and ICOS (HR:0.6, p=0.047) in tumour compartments were associated with improved overall survival (OS).

## Results

### Region of interest (ROI) selection

Ninety-six ROIs in total were selected that were representative of 45 tumour, 32 TME and 19 histologically normal adjacent tissues from the cohort of patients described in **Table 2**. Images of H&E stained cores were demarcated by a pathologist and were utilised alongside Nanostring**™** immunofluorescent staining for morphology markers PanCK, CD45, CD3 and DAPI to draw ROIs indicative of tumour (CK+) or TME (CK-/CD3+). **Figure 1** provides an example of this strategy where tumour and TME ROIs were able to be identified within the same tumour core. Of all samples collected, comparisons from the same patient could be made between 8 TME-NAT pairs, 14 NAT-tumour pairs and 18 tumour-TME pairs. **Figure 2** provides an overview of the tumour and immune ROI selection and representative expression profiles for a number of associated markers.

**Figure 1.**
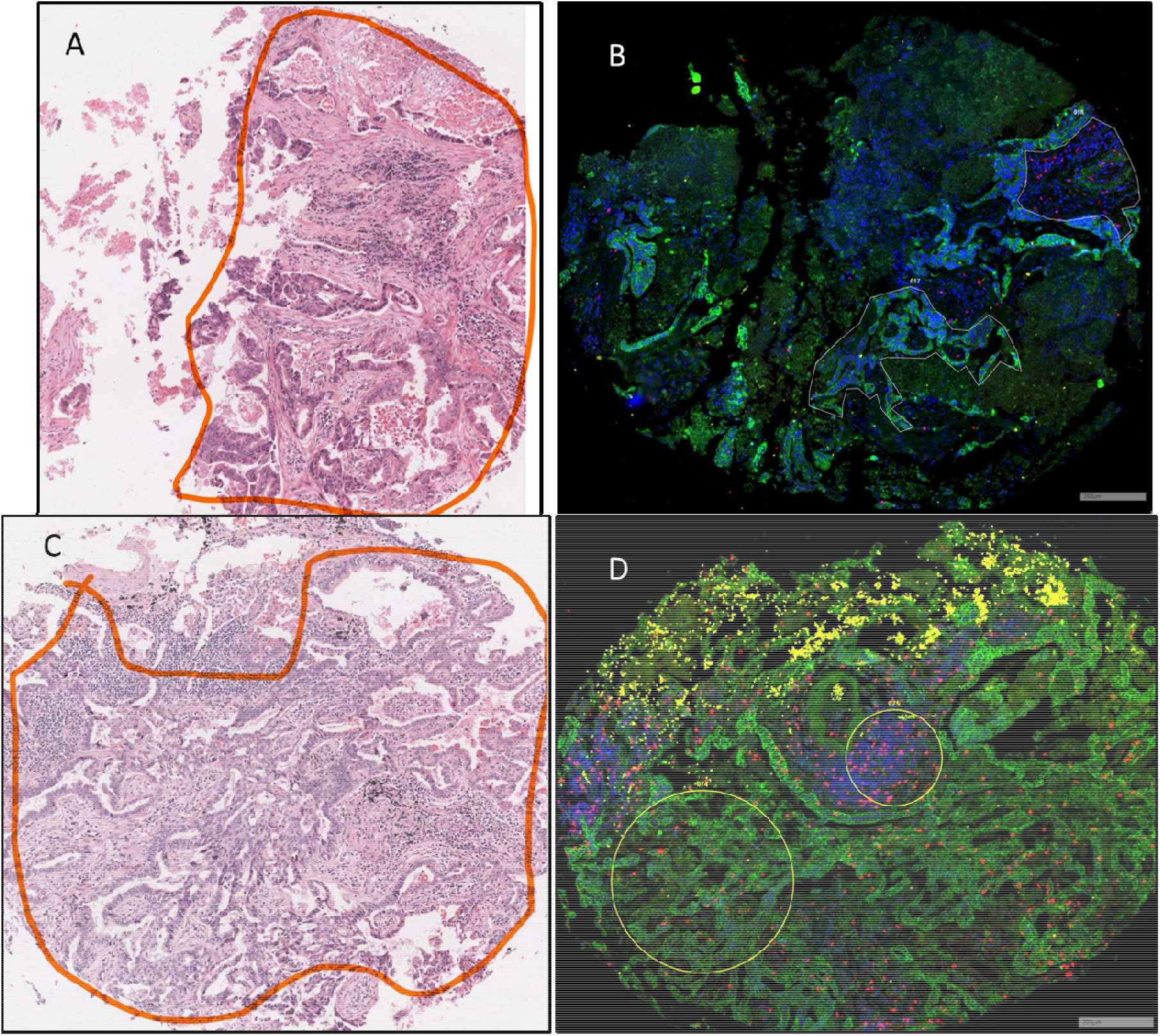
Representative H&E and immunofluorescent images of TMA cores. (A,C) Tumour regions of cores were demarcated by pathologist, and; B,D) Corresponding ROIs were captured for DSP analysis based on immunofluorescent staining for PanCK (Green), CD3 (Red), CD45 (Yellow) and DAPI (Blue). Tumour ROI (lower left) and TME ROI (upper right) were manually drawn in (B), and circular tumour ROI (lower left) and TME ROI (upper right) were used in (D). Scale bar not available for H&E images as images obtained from commercial supplier and are representative only. Scale bar = 200µm.

**Figure 2.**
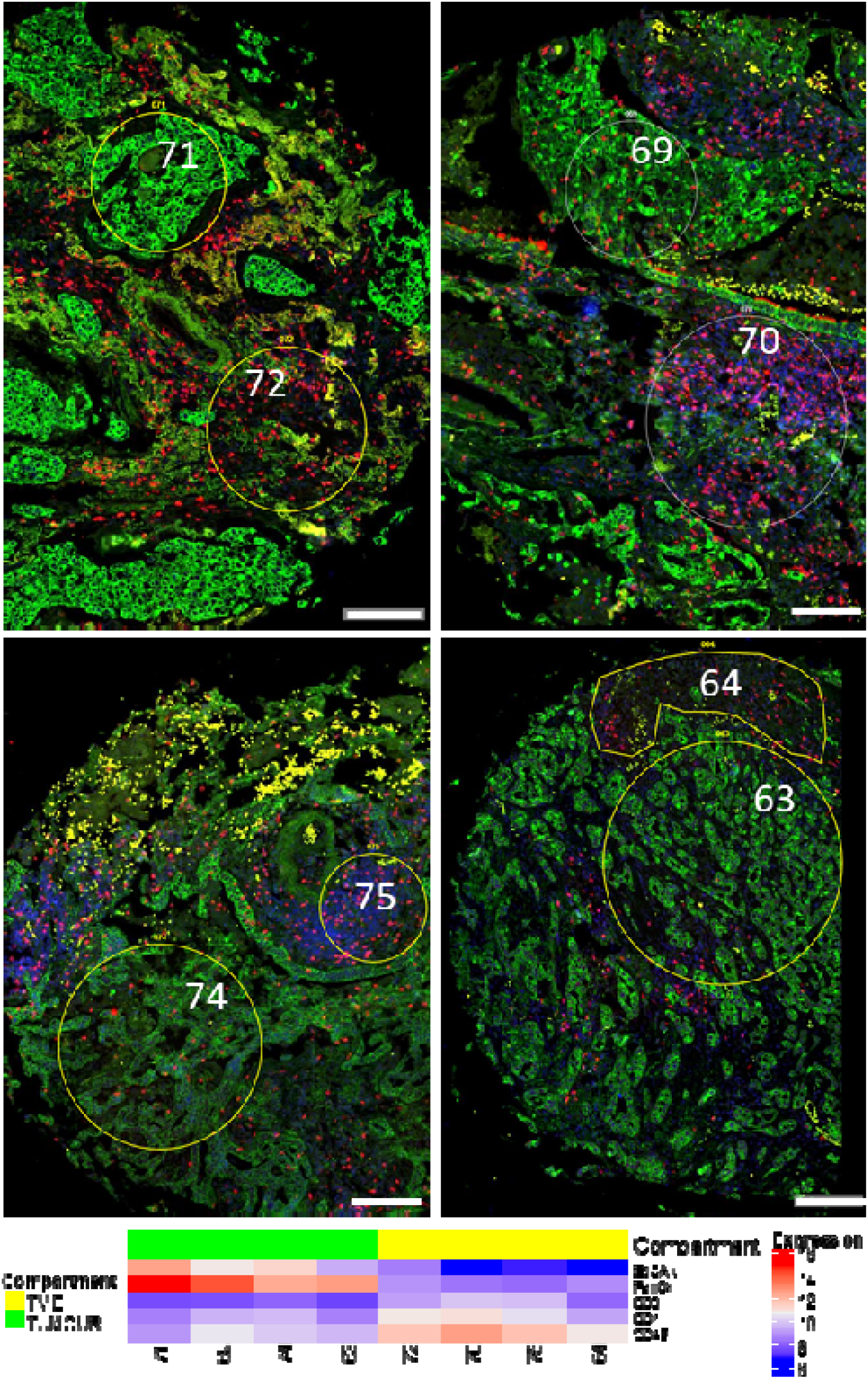
Tumour and immune ROIs exhibit distinct tumour/leukocyte marker expression. PanCK (Green), CD3 (Red), CD45 (Yellow) and DAPI (Blue). Representative paired tumour and immune ROIs are shown with corresponding Log2 expression of EpCAM, PanCK, CD3, CD4 and CD45. ROIs 71, 69, 74, 63 represent tumour regions, while 72, 69, 74, 63 are respective immune regions from corresponding patients. Scale bar = 200µm.

### Data quality control

Quality control was performed within GeoMx™ DSP analysis suite, to ensure Ncounter® quantification of probes was within specification. Probe counts per ROI were inspected to ensure comparable ranges of signal. ROIs generated median counts within range of 10^2^ and 10^3^, with observably lower median counts for ROIs 13, 67, and 96 (**Supplementary Figure S1**). Probe counts were then inspected within TME, Tumour, and NAT, as targets were expected to vary by respective tissue compartment (e.g. immune targets in TME vs tumour). Robust counts were observed for abundant targets, including Histone H3, SMA, S6, GAPDH, fibronectin, cytokeratin, CD44, CD68, β-2-microglobulin (B2M), HLA-DR, CD45 and B7-H3 (beyond axis range in **Supplementary Figure S2**), however the remaining probes exhibited raw counts below 200, including negative control IgG probes which possessed counts between 50 to 150 (**Supplementary Figure S2**). Rabbit (Rb) IgG exhibited similar counts between tumour and TME compartments, while mouse (Ms) IgG showed higher counts for tumour then TME, suggesting that background correction and normalisation would be important considerations in legitimate quantification of lowly expressed targets. Target signal relative to Ms and Rb IgG control probes was therefore evaluated to identify probes from which data should be considered with caution. Probes shown in **Figure 3** whose median signal relative to IgG was less than 1 (pink region) were thus followed with caution, and 31 of 55 probes above CD25 in **Figure 3** below were considered robust for further analysis.

**Figure 3.**
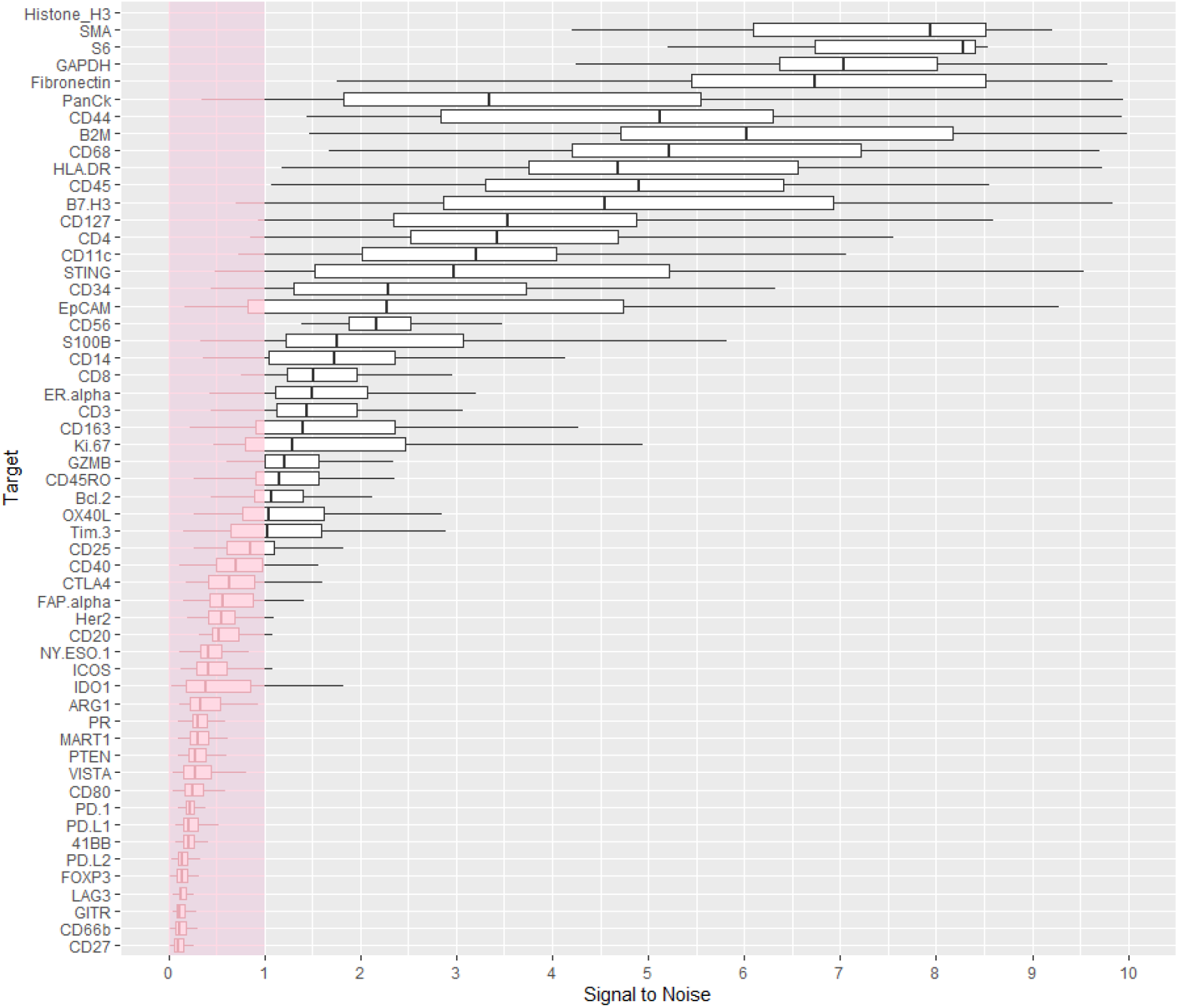
Probe counts relative to Rb and Ms IgG controls. Counts of each probe were normalised to mean counts of Rb and Ms IgG within each ROI. The mean of these normalised values per probe was plotted to evaluate the robustness of target protein signal to non-specific background.

### Data Normalisation

The method of normalisation between ROIs was assessed by examining correlations between Histone H3, S6, GAPDH and IgG background control probes, under the assumption that normalisers should correlate between ROIs and be unrelated to underlying biology. Housekeeping proteins GAPDH, Histone H3 and S6 were plotted to determine which pairs best correlated across ROIs (**Figure 4 A-C**). Histone H3 and S6 exhibited the strongest Pearson correlation coefficient (R=0.7) (**Figure 4C**) and were thus examined further for correlation to IgG background to confirm independence from tissue biology. Ms and Rb IgG strongly correlated with each other across ROIs (R=0.92) (**Figure 4D**), and the means of these IgG counts showed strong correlation with means of Histone H3 and S6 housekeeping controls (R=0.91), indicating that both IgG controls and Histone H3 and S6 were unrelated to underlying biology and could act as appropriate normalisers across ROIs (**Figure 4E**).

**Figure 4.**
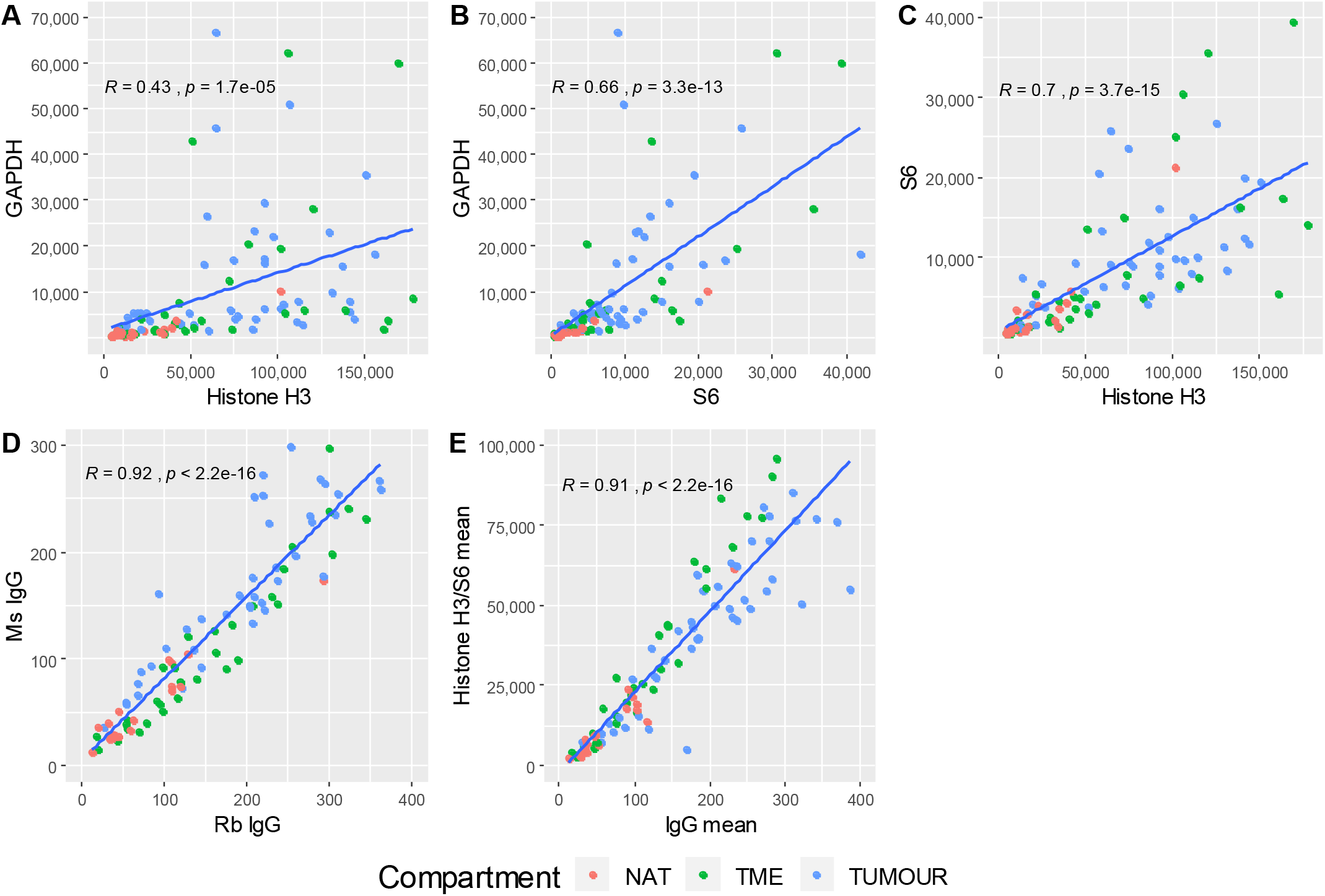
Assessment of housekeeping proteins and IgG as normalisers. Raw counts (A-D) and the means (E) of GAPDH, Histone H3, S6, Ms/Rb IgG were compared pairwise and linear regression performed to determine the Pearson correlation coefficient (R) between putative housekeepers. A) GAPDH vs Histone H3, R=0.43; B) GAPDH vs S6, R=0.66; C) Histone H3 vs S6, R=0.7; D) Ms IgG vs Rb IgG, R=0.92; E) mean of Ms/Rb IgG vs mean of Histone H3/S6, R=0.91. NAT: Normal adjacent tissue; TME: Tumour microenvironment; Tumour: Tumour region.

In addition to the normalisation by IgG and traditional housekeeping members, ROI area and nuclei count were inspected for their utility to normalise DSP data. (**Figure 5 A-C)** illustrates the relationship that ROI area possessed with Histone H3/S6, IgG and nuclei. A number of ROIs varied by area, however some possessed maximum sized geometry, and these ROIs varied significantly in their relationship with other normalisation parameters, indicating that ROI area was not a useful normalisation method in this experiment. Similarly, nuclei counts were evaluated relative to IgG and Histone H3/S6 means (**Figure 5 D-E**), where some trend was evident, but lacked the robustness of either IgG or Histone H3/S6 normalisation. Histone H3/S6 means were therefore utilised for normalisation, and henceforth comparative quantification of probes.

**Figure 5.**
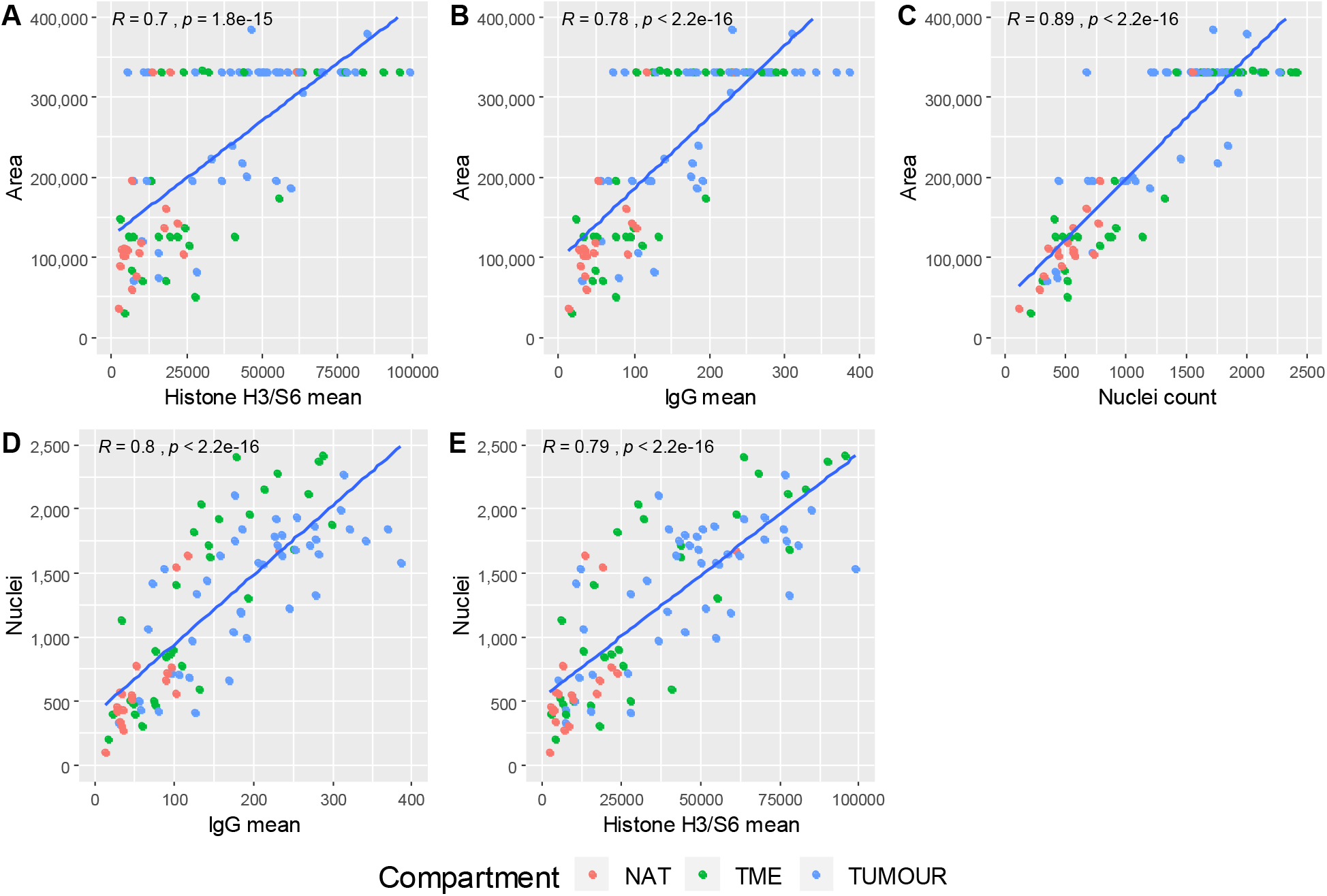
Assessment of ROI area and Nuclei as normalisers. (A) ROI area was plotted against Histone H3/S6 means, R= 0.7; (B) IgG means, R= 0.78; (C) Nuclei counts, R=0.89. Nuclei counts were then evaluated against (D) IgG means, R= 0.8; and (E) Histone H3/S6 means, R= 0.79. Some ROIs contained the maximum area (horizontal dots in A-C) and exhibited significant variance in secondary parameter, indicating area was not a suitable normalisation method. Nuclei counts (D-E) demonstrated a trend with secondary parameter, however correlation was not as significant as that observed for IgG or Histone H3/S6. NAT: Normal adjacent tissue; TME: Tumour microenvironment; Tumour: Tumour region.

### Data analysis

Hierarchal clustering by the Ward D2 method ^14^ was first used to explore normalised data, however expression appeared to vary significantly within classes of compartments, such that clear distinction between NAT, tumour and TME was not evident (**Figure 6**). K-means clustering to further group ROIs into classes showed most NATs to group together (left **Figure 6**), characterised by higher expression of most genes except for PanCK, EpCAM and Ki-67. Another class consisting of both TME and tumour (middle **Figure 6**) was characterised by lower expression of most proteins with some ROIs expressing high levels of Ki-67 and EpCAM, whereas a third class was characterised by relatively heterogenous expression of all proteins (right **Figure 6**).

**Figure 6.**
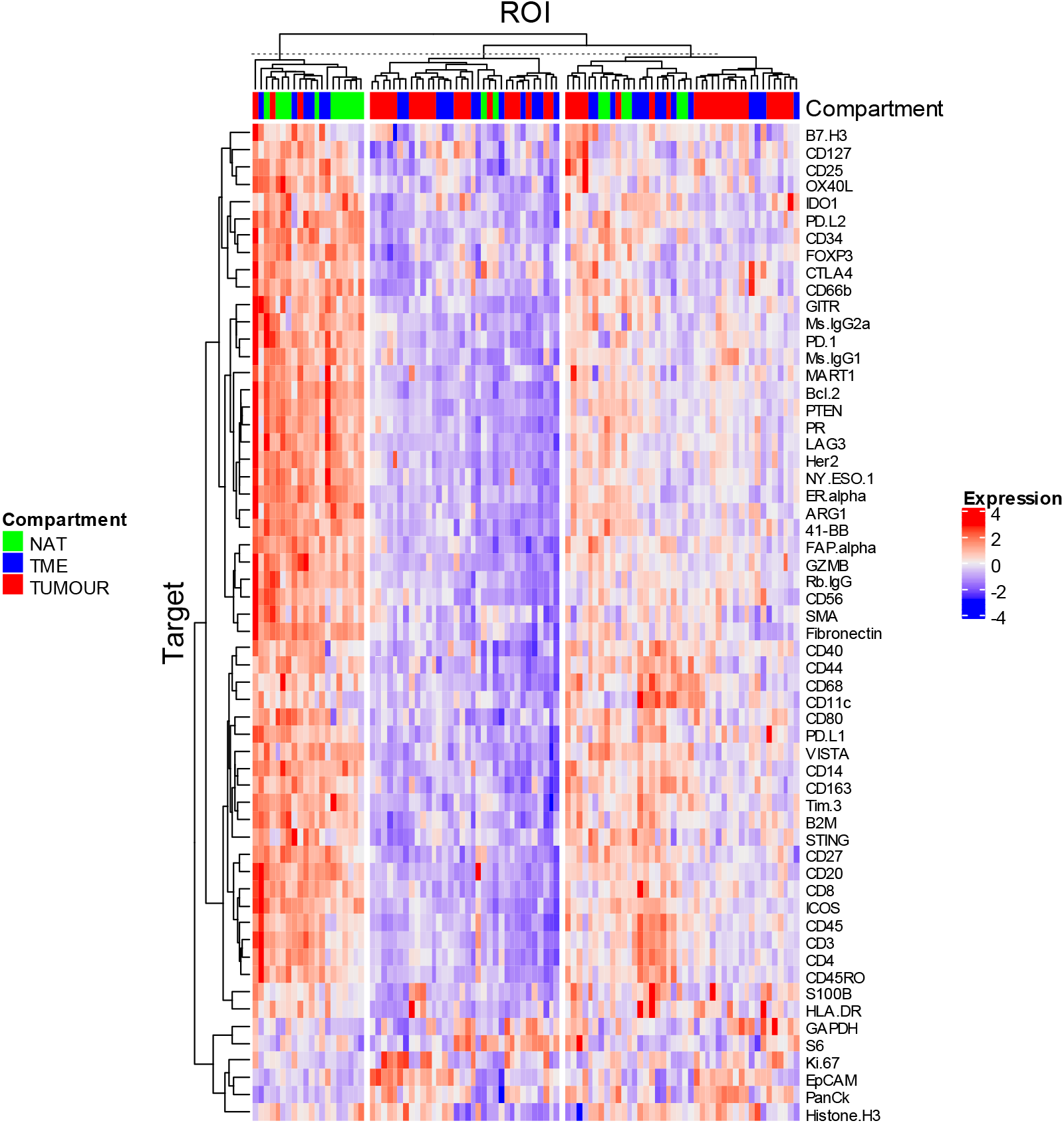
Clustered heatmap of relative expression of proteins per ROI. Ward D2 clustering was applied, followed by K-means clustering to delineate differences between expression profiles amongst compartments. NAT: Normal adjacent tissue; TME: Tumour microenvironment; Tumour: Tumour region.

Global correlation matrices for target protein expression within TME (**Supplementary Figure S3**) and tumour (**Supplementary Figure S4**) indicated a large number of significant (p ≤ 0.001) positive correlations.

Differential protein expression between patient matched compartments was then evaluated (**Figure 7**). Interestingly, matched TME and NAT (n=8) did not exhibit significant differences (**Figure 7A**). Matched TME-tumour pairs (n=18) identified the enrichment of CD27, CD3, CD4, CD44, CD45, CD45RO, CD68, CD163, VISTA within TME, while tumour regions were enriched in Ki-67, EpCAM and cytokeratin (**Figure 7B**). As expected, most proteins were differentially expressed in NAT relative to tumour ROIs (n=14) (**Figure 7C**).

**Figure 7.**
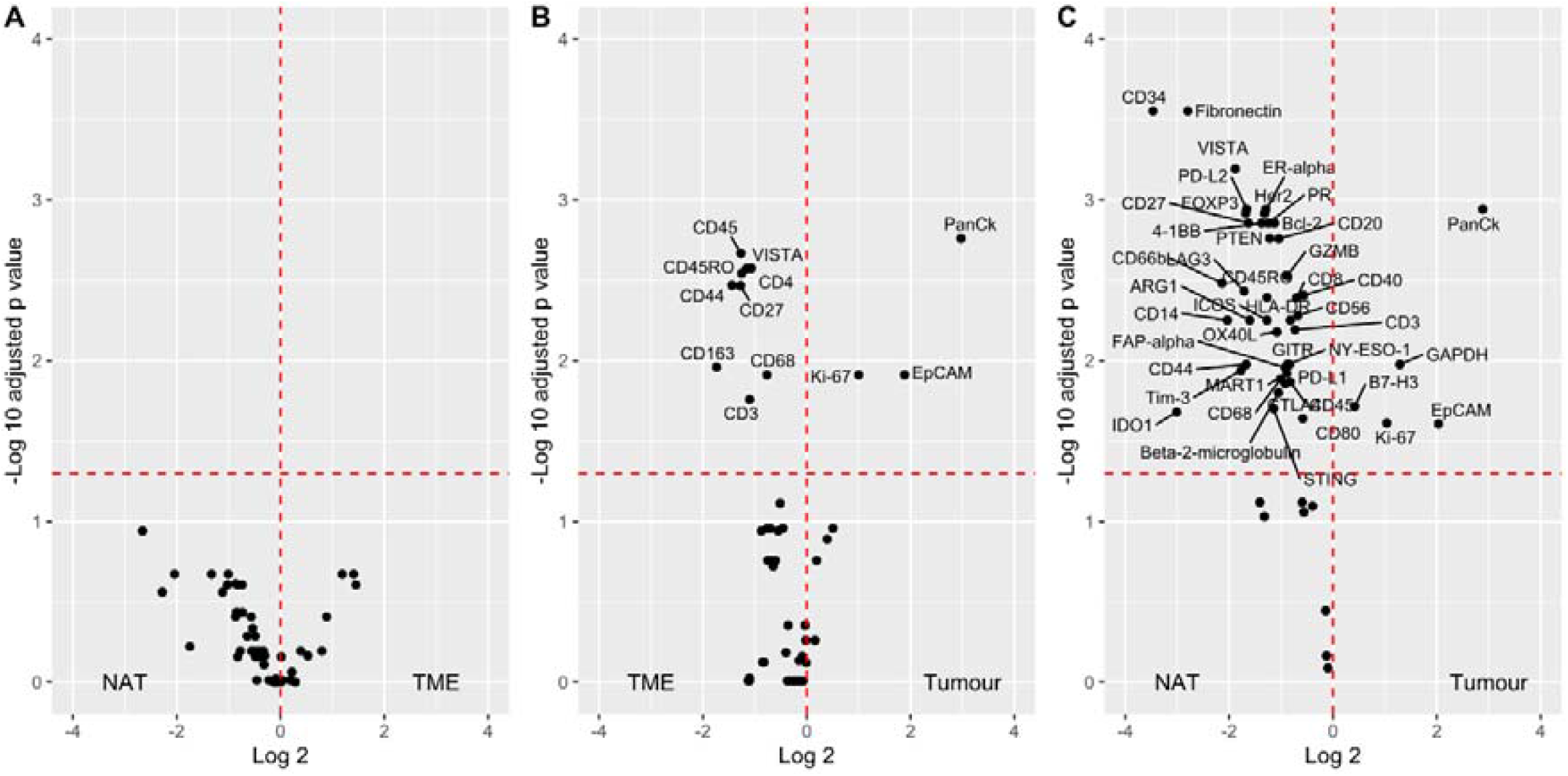
Differential expression of proteins between patient paired tissue compartments. Paired t-tests with Benjamini-Hochberg correction were performed between compartments, and p values adjusted for multiple testing were used to identify significantly differentially expressed proteins. A) NAT-TME (n=8), B) TME-tumour (n=18), C) NAT-tumour (n=14) NAT: Normal adjacent tissue; TME: Tumour microenvironment; Tumour: Tumour region.

When incorporating all samples irrespective of patient pairing, several proteins appeared to be downregulated in TME relative to NAT, including CD34, fibronectin, IDO1, LAG3, ARG1 and PTEN (**Figure 8A**). TME-tumour comparisons remained similar to the paired data, where CD3, CD45RO, VISTA and CD163 were enriched in TME relative to tumour. Univariate assessment of the association between protein expression and survival was explored through Cox proportional hazards regression. Interestingly, expression data from immune ROIs indicated that the presence of EpCAM and cytokeratin were associated with better patient OS (**Figure 9A**), while the presence of CD34, CD3 and ICOS in tumour ROIs were associated with better patient OS. The number of samples did not permit further multivariate analysis and statistical modelling of signatures indicative of patient outcome.

**Figure 8.**
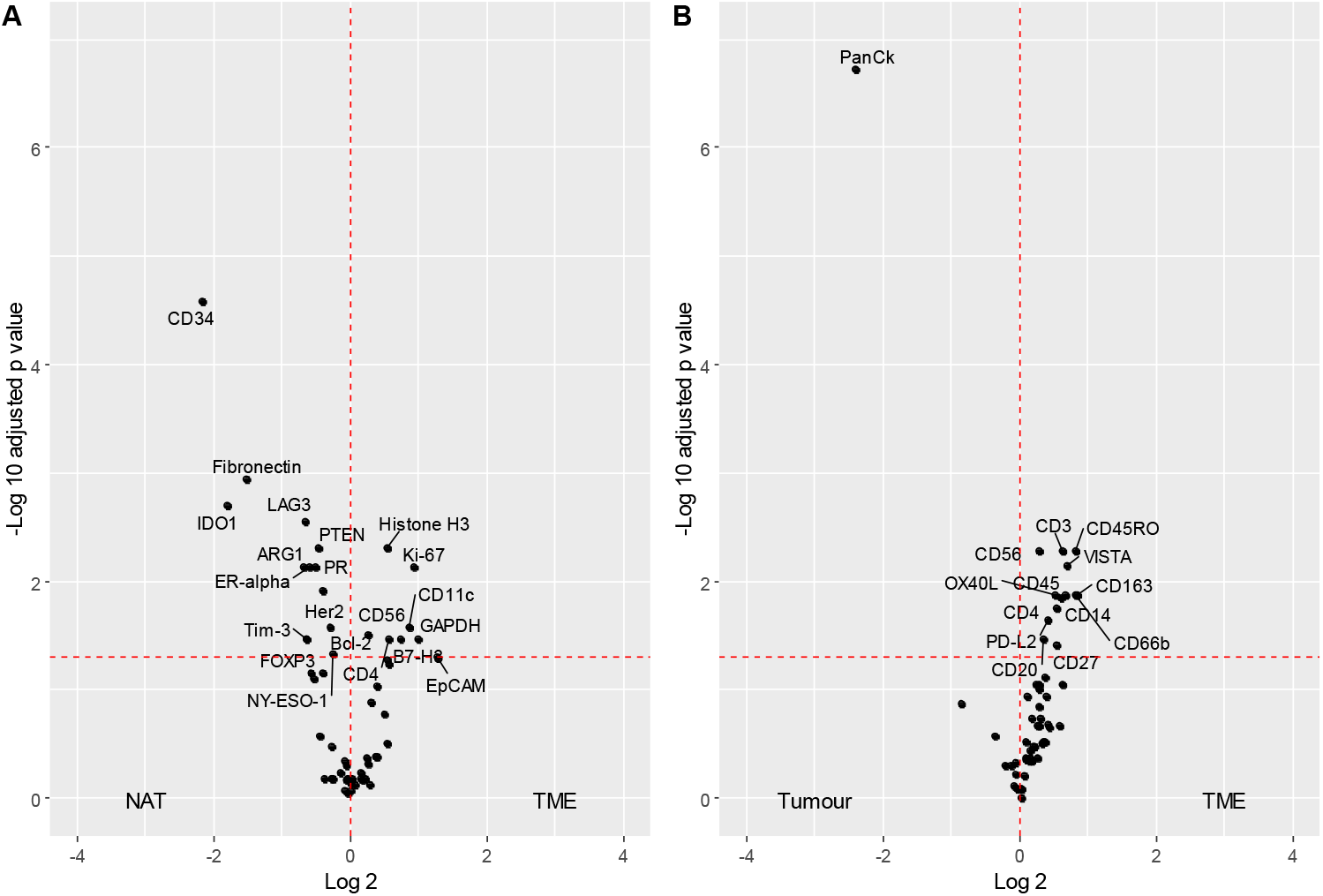
Differential expression of proteins between unmatched compartments. Mann Whitney tests with Benjamini-Hochberg correction were performed between compartments, and p values adjusted for multiple testing were used to identify significantly differentially expressed proteins. A) NAT (n=19) vs TME (n=32); B) Tumour (n=45) vs TME. NAT: Normal adjacent tissue; TME: Tumour microenvironment; Tumour: Tumour region

**Figure 9.**
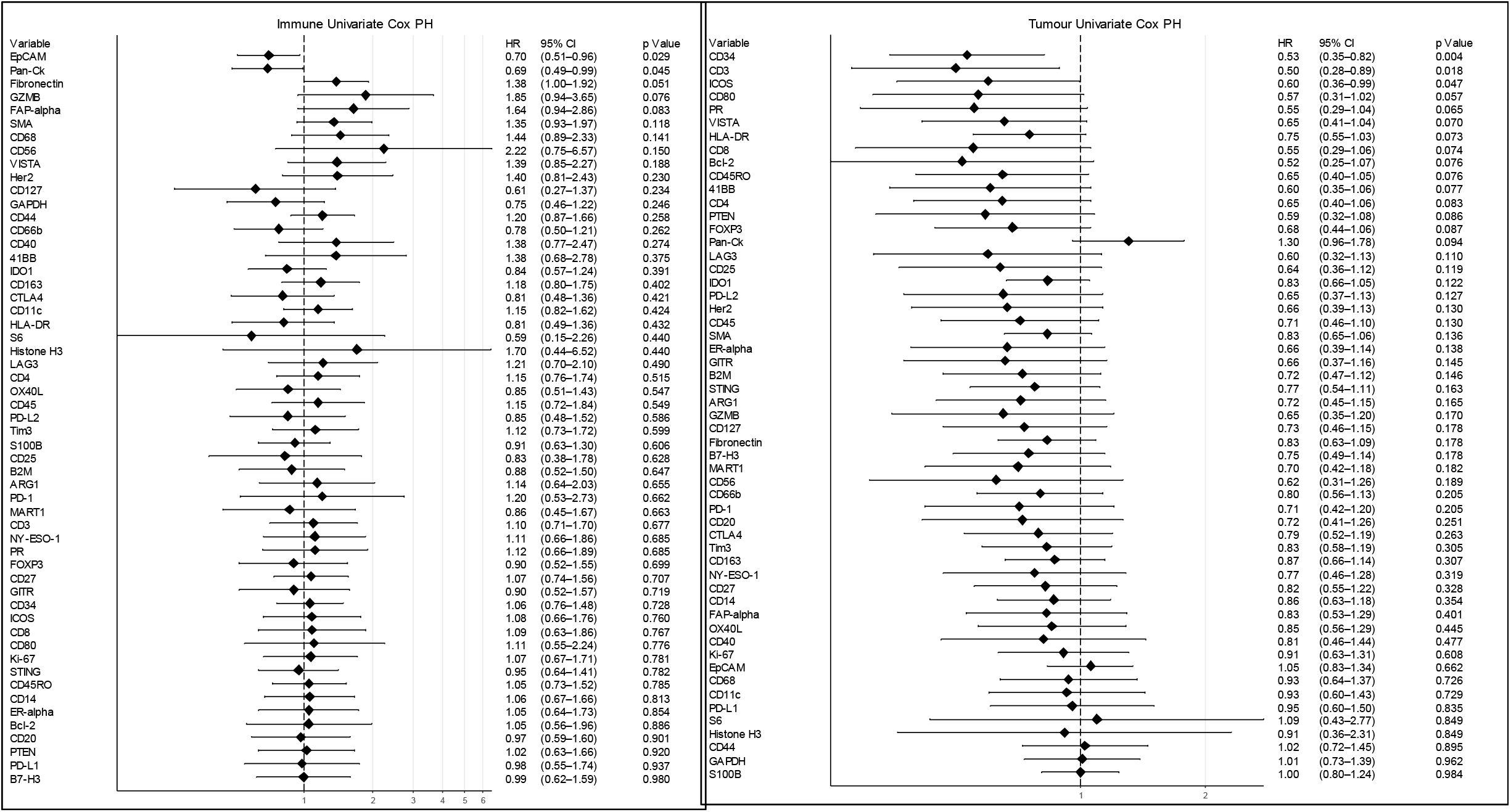
Cox proportional hazards of compartment specific protein expression ranked by association with overall survival. Log2 protein expression was modelled against follow-up time for: A) TME; B) Tumour. Hazard ratio (HR) <1 = associated with better patient outcome, HR>1 associated with poorer patient outcome.

## Discussion

The Nanostring GeoMx™ DSP platform ^13,15^ offers a novel solution for high plex, digital quantification of protein and mRNA from fixed and fresh frozen tissues with spatial resolution. It has been recently applied to triple negative breast cancer (TNBC) ^16^, NSCLC ^17^ and Melanoma ^18^, however the implementation, and interpretation of such high-plex discovery is still in its infancy. The application of such technologies to large numbers of patient samples in TMA format potentially provides unparalleled insight into spatial cell types, biomarkers and the interactions that may underlie the disease biology. In this study, we quantified proteins across the current DSP immune cell profiling, IO drug target, immune activation status, immune cell typing, and pan-tumour protein modules to understand the presence of these markers in tumour, tumour microenvironment and histologically normal adjacent tissue compartments.

In conventional IHC and multiplex IHC, information can be obtained from the entirety of sections or TMA cores, giving a global perspective of marker expression, and allowing post-hoc segmentation to inform on distribution. The DSP approach differs in that while visualisation markers may inform on tumour/non-tumour regions and areas of immune cell infiltrate, ROIs are limited to a maximum of 600 µm geometric shapes. In this study, circular ROIs and several custom drawn ROIs were used, meaning that ‘tumour’ ROIs innately contained immune infiltrate and that ‘immune’ ROIs needed to be completely separate from tumour in order to be defined, and may represent tumour-adjacent ‘stromal’ immune infiltrate rather than an activated ‘tumour microenvironment’ immune infiltrate. The DSP platform does allow for ‘masking’ or ‘compartmentalization’ within ROIs, enabling signal to be obtained directly from tumour cells and from the immediate stromal space into which they have proliferated, at µm resolution ^17^, however this approach was not used in this study, and is a salient point to be considered for future analyses using the platform.

Here, we also demonstrate that there is a need to empirically determine the method of normalisation and identify probes which lack robust signal to noise. We demonstrated that both IgG background control probes and Histone H3, S6 housekeeping probes significantly correlated across ROIs, while area and nuclei varied significantly and were thus less reliable to normalise data for quantification. Existing studies that have used area as a normaliser ^17^, also utilised signal-to-noise ratio cut-off >3, suggesting that our particular TMA may have exhibited disproportionately high background, or low overall signal, as a significant number of probes were within range of IgG control probes. It is perhaps for this reason that many significant correlations were observed within compartments for markers that possessed low signal to noise, and these observations require additional validation.

Nevertheless, it was notable that when patient pairing was applied to a limited sample size, NAT was indistinguishable from TME by differential analysis. A clear distinction between matched tumour and TME was evident however, and was indicated by the increased presence of several key markers within the TME. Such markers included CD44, CD45, T cell lineage (CD3, CD4), memory T cell (CD45RO), monocyte/macrophage lineage (CD163, CD163) and costimulatory immune checkpoints (CD27, VISTA). When incorporating all samples irrespective of patient matching, immunosuppressive molecules LAG3 19 and IDO1 20 were, perhaps counter-intuitively, significantly depleted in the TME relative to NAT, indicating the requirement for patient matching to make meaningful comparisons.

Nevertheless, the sheer scale of high-plex analyses appropriately applied to large numbers of cases through TMAs is an incredibly powerful tool for spatial biology. The DSP protein modules include key markers that describe multiple immune cell types, immune checkpoints and experimental targets that enable a more comprehensive understanding of the immunological parameters that influence patient outcome. While overall survival was the only clinical endpoint investigated in this study, the emergence of patient cohorts treated with immunotherapies mean that such assays may be used to track patient progression and outcomes, and indicate potential biomarkers for patients most likely to respond to these therapies. Despite some limitations in the absolute definition of tumour and TME compartments in our study, we were able to identify that the presence of CD3, CD34 and ICOS expressing cells in tumour compartments were associated with better patient OS significantly.

It is interesting to note that enrichment of naïve T cell marker, CD3, was significantly associated with improved OS in this study independently of additional activation markers such as CD4 and CD8. Several markers significantly correlated with CD3 expression in the tumour compartment, including CD40, CD44, CD14, B2M, Tim-3, CD8, CD45RO and ICOS, potentially implicating other cell lineages in immune associated anti-tumour activity. Of note is the correlation between CD3 and ICOS, both of which were independently prognostic within tumour compartments, highlighting the power of such multiplex discovery, and the requirement for orthogonal validation.

In summary, the application of such novel platforms to provide comprehensive snapshots of clinical material enables an unprecedented insight into the molecular phenotypes that may be indicative of response to emerging therapies, and ultimately, patient outcome. By combining such high-plex approaches with TMAs and orthogonal validation through multispectral IHC, a new field of biomarker discovery is developing that offers to change the way clinical pathology is performed.

## Methods

### Tissue Microarray

NSCLC TMA (HLugA180Su03) containing 92 cases with concordant histologically normal adjacent tissue was obtained from US Biomax, Inc. (Rockville, MD, USA), including associated clinical information. H&E images were demarcated by a pathologist for tumour and non-tumour regions in each core

### Nanostring GeoMX™ Digital Spatial Profiler: tissue microarray

The slides were profiled using Technology Access Program (TAP) by Nanostring™ Technologies (Seattle, WA, USA). In brief, immunofluorescent staining was performed on the TMA with tissue morphology markers (PanCK, CD3, CD45 and DAPI) in parallel with DNA barcoded antibodies within the immune cell profiling, IO drug target, immune activation status, immune cell typing, and pan-tumour protein panels as shown in **Table 1**. Geometric (circular) and custom ROIs were selected based on visualisation markers to generate tumour (PanCK+) and TME (PanCK-) areas from which barcodes were liberated by UV light by the GeoMx™ DSP instrument, hybridised, and counted on the Ncounter® system.

**Table 1.** Target proteins within Nanostring™ DSP modules

| Controls | Immune Cell Profiling | IO Drug Target | Immune Activation Status | Immune Cell Typing | Pan-Tumour Module |
| --- | --- | --- | --- | --- | --- |
| Rb IgG | PD-1 | 4-1BB | CD127 | CD45RO | MART1 |
| Ms IgG1 | CD68 | LAG3 | CD25 | FOXP3 | NY-ESO-1 |
| Ms IgG2a | HLA-DR | OX40L | CD80 | CD34 | S100B |
| Histone H3 | Ki-67 | Tim-3 | ICOS | CD66b | Bcl-2 |
| S6 | Beta-2M | VISTA | PD-L2 | FAP-alpha | EpCAM |
| GAPDH | CD11c | ARG1 | CD40 | CD14 | Her2 |
|  | CD20 | B7-H3 | CD44 | CD163 | PTEN |
|  | CD3 | IDO1 | CD27 |  | ER-alpha |
|  | CD4 | STING |  |  | PR |
|  | CD45 | GITR |  |  |  |
|  | CD56 |  |  |  |  |
|  | CD8 |  |  |  |  |
|  | CTLA4 |  |  |  |  |
|  | GZMB |  |  |  |  |
|  | PD-L1 |  |  |  |  |
|  | PanCk |  |  |  |  |
|  | SMA |  |  |  |  |
|  | Fibronectin |  |  |  |  |

### Nanostring GeoMX™ Digital Spatial Profiler: data analysis

Patient data presented in Table 2 was generated in R studio ^21^ using package “gtsummary” ^22^. Remote access to the GeoMx™ DSP analysis suite (GEOMX-0069) allowed inspection, Quality Control (QC), normalisation, and differential expression to be performed. Briefly, each ROI was tagged with metadata for its compartment and patient pairing to allow pairwise comparisons. Raw data was exported and plotted in R using “ggplot2” ^23^ for raw counts, signal relative to IgG controls, and evaluation of Pearson correlation coefficients (R) between normalisation parameters using “ggpubr” package ^24^. Normalisation using Histone H3 and S6 proteins was performed in GeoMx™ DSP analysis suite. Differential expression between paired compartments was evaluated by paired T-tests with Benjamini-Hochberg correction while differential expression between unpaired compartments performed by Mann Whitney test with Benjamini-Hochberg correction and results plotted in R studio using “ggplot2”. Relative expression data was exported from GeoMx™ DSP analysis suite and hierarchical clustering performed using R package “complexHeatmap” ^25^ and univariate Cox proportional hazards regression performed using “survivalAnalysis” ^26^ package.

**Table 2.** Patient characteristics of the TMA cohort by DSP tissue compartment.

| Characteristic | Overall, N = 96 | NAT, N = 19<sup>1</sup> | TME, N = 32<sup>1</sup> | TUMOUR, N = 45<sup>1</sup> | p-value<sup>2</sup> |
| --- | --- | --- | --- | --- | --- |
| **Age** | 62 (54, 69) | 66 (56, 69) | 60 (54, 67) | 62 (54, 71) | 0.6 |
| **Sex** |  |  |  |  | 0.7 |
| F | 42 (44%) | 9 (47%) | 12 (38%) | 21 (47%) |  |
| M | 54 (56%) | 10 (53%) | 20 (62%) | 24 (53%) |  |
| **T** |  |  |  |  |  |
| 0 | 19 (20%) | 19 (100%) | 0 (0%) | 0 (0%) |  |
| 1 | 24 (25%) | 0 (0%) | 11 (34%) | 13 (29%) |  |
| 2 | 34 (35%) | 0 (0%) | 15 (47%) | 19 (42%) |  |
| 3 | 12 (12%) | 0 (0%) | 4 (12%) | 8 (18%) |  |
| 4 | 7 (7.3%) | 0 (0%) | 2 (6.2%) | 5 (11%) |  |
| **N** |  |  |  |  | 0.2 |
| 0 | 78 (81%) | 19 (100%) | 23 (72%) | 36 (80%) |  |
| 1 | 13 (14%) | 0 (0%) | 6 (19%) | 7 (16%) |  |
| 2 | 3 (3.1%) | 0 (0%) | 2 (6.2%) | 1 (2.2%) |  |
| 3 | 2 (2.1%) | 0 (0%) | 1 (3.1%) | 1 (2.2%) |  |
| <sup>1</sup> Statistics presented: median (IQR); n (%) |  |  |  |  |  |
| <sup>2</sup> Statistical tests performed: Kruskal-Wallis test; chi-square test of independence; Fisher's exact test |  |  |  |  |  |

## Author Contributions

Idea/Concept: AK, KOB; Experimentation: JM, AK, CO, TT; Data analysis: JM, AK, KOB, DR, MEW; Critical review and writing of manuscript: all authors.

## Data Availability

All data from this study will be deposited in a publicly archived database.

## Acknowledgements

This study was funded by the Princess Alexandra Hospital Foundation grant for KOB. AK is supported by an NHMRC ECF Fellowship (APP1157741) and Cure Cancer (APP1182179). The authors are grateful for the assistance by Nanostring Seattle (Liuliu Pan) and Singapore (Caroline Chan).

## Supplementary Figures

**Figure S1.**
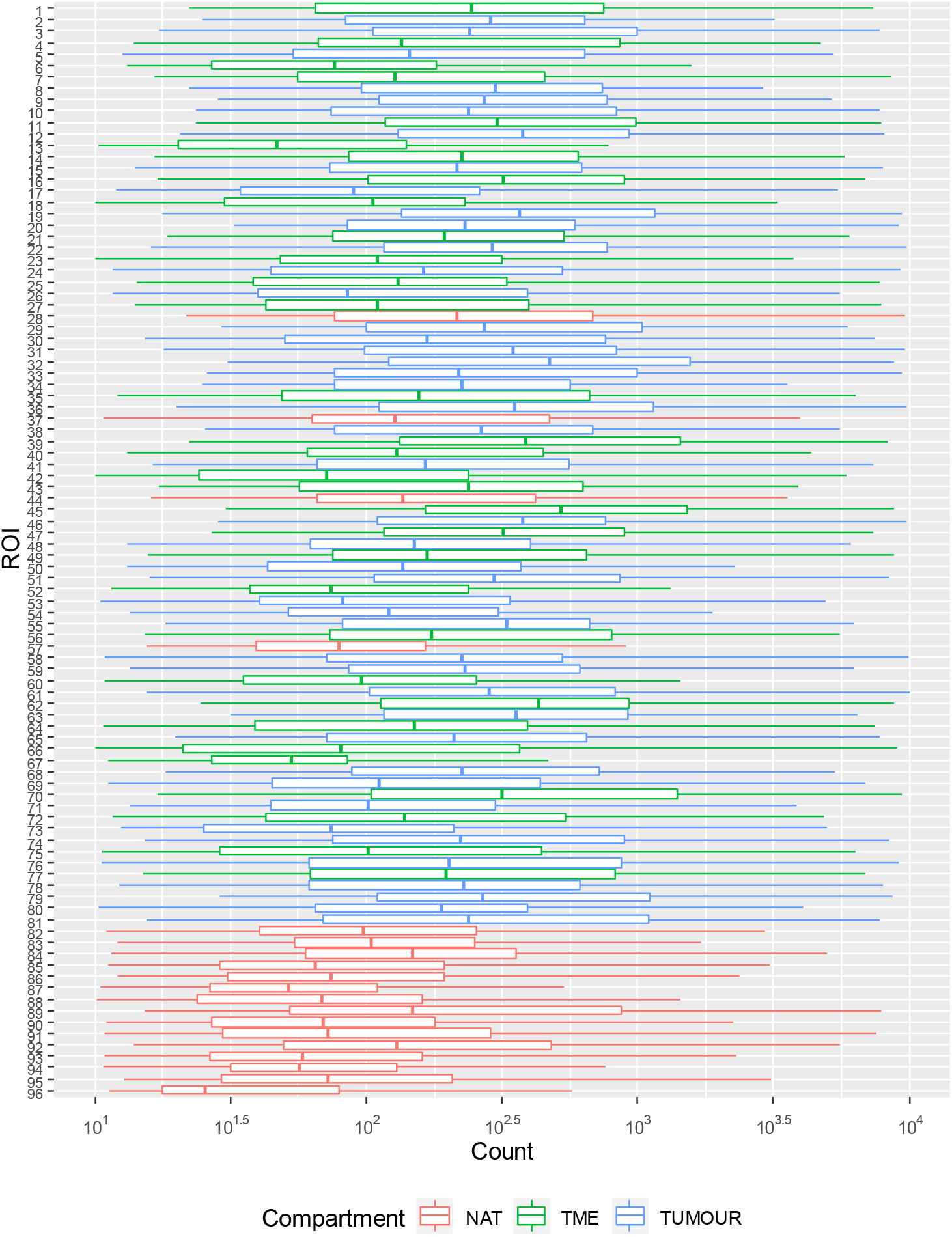
Range of probe counts from ROI-1 (Top) to ROI-96 (Bottom). Raw counts for all probes per ROI were plotted to confirm comparable ranges of signal between ROIs. NAT: Normal adjacent tissue; TME: Tumour microenvironment; Tumour: Tumour region.

**Figure S2.**
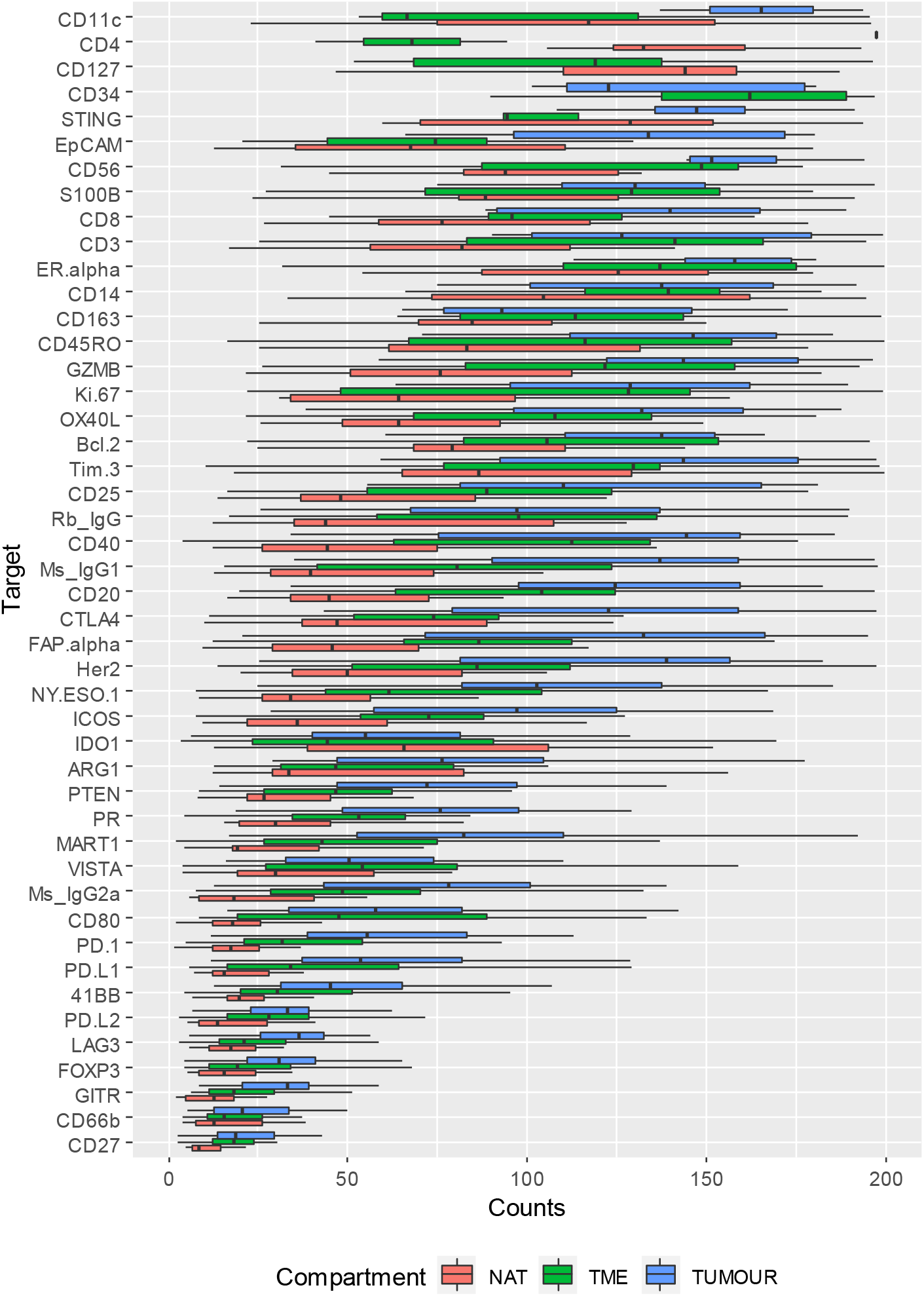
Probe counts per compartment from most abundant (Top) to least abundant (Bottom). Raw counts for each probe were plotted to examine compartment specific expression and quantifiable range of lowly expressed targets. Counts for highly abundant Histone H3, SMA, S6, GAPDH, fibronectin, cytokeratin, CD44, CD68, β-2-microglobulin (B2M), HLA-DR, CD45 and B7-H3 excluded here. NAT: Normal adjacent tissue; TME: Tumour microenvironment; Tumour: Tumour region.

**Figure S3.**
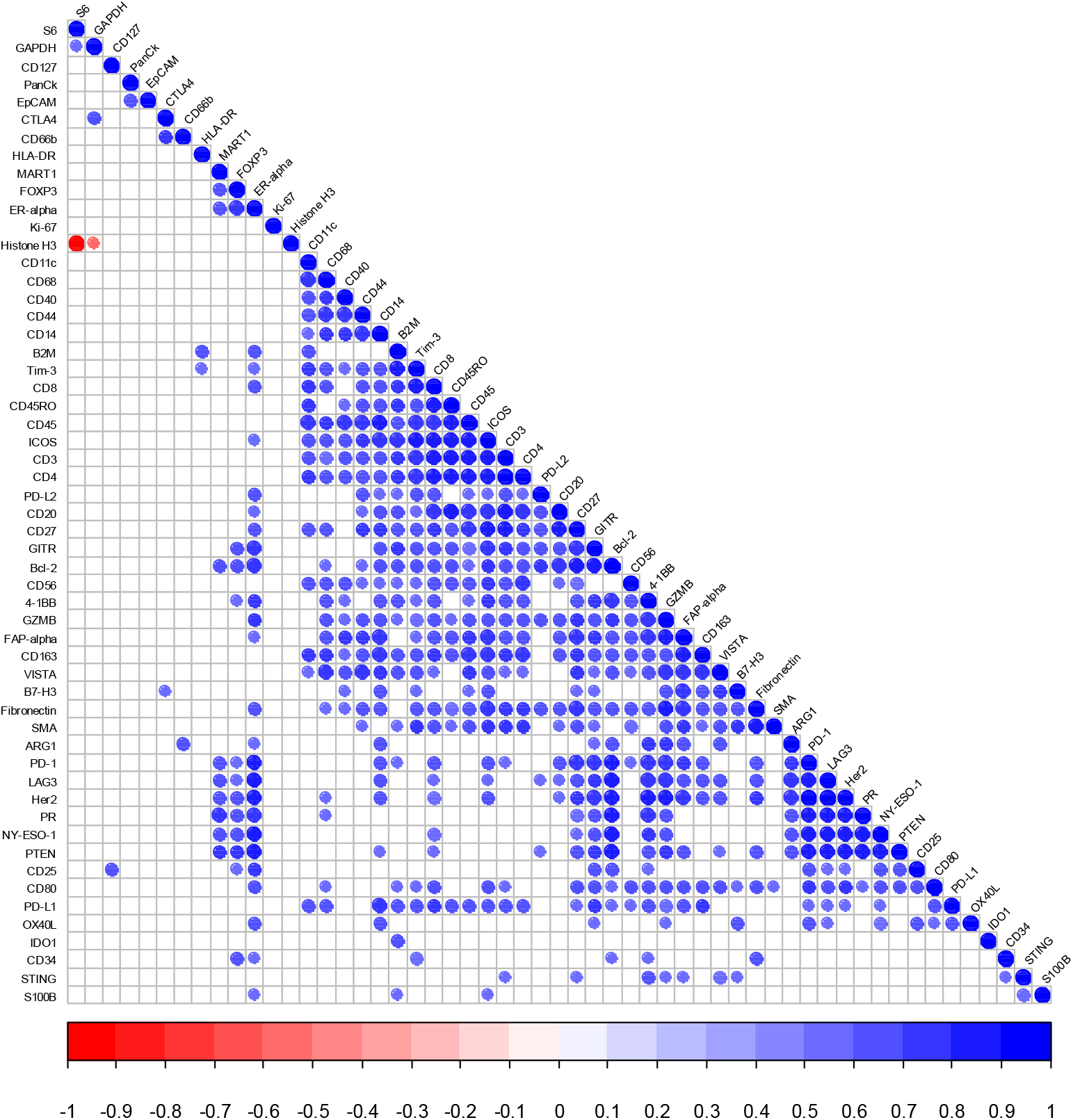
Pearson correlation matrix of proteins within TME ROIs. Log2 protein expression was used to evaluate Pearson correlations between proteins. Only correlations with p value <0.001 are shown. Blue= positive correlation, Red= negative correlation

**Figure S4.**
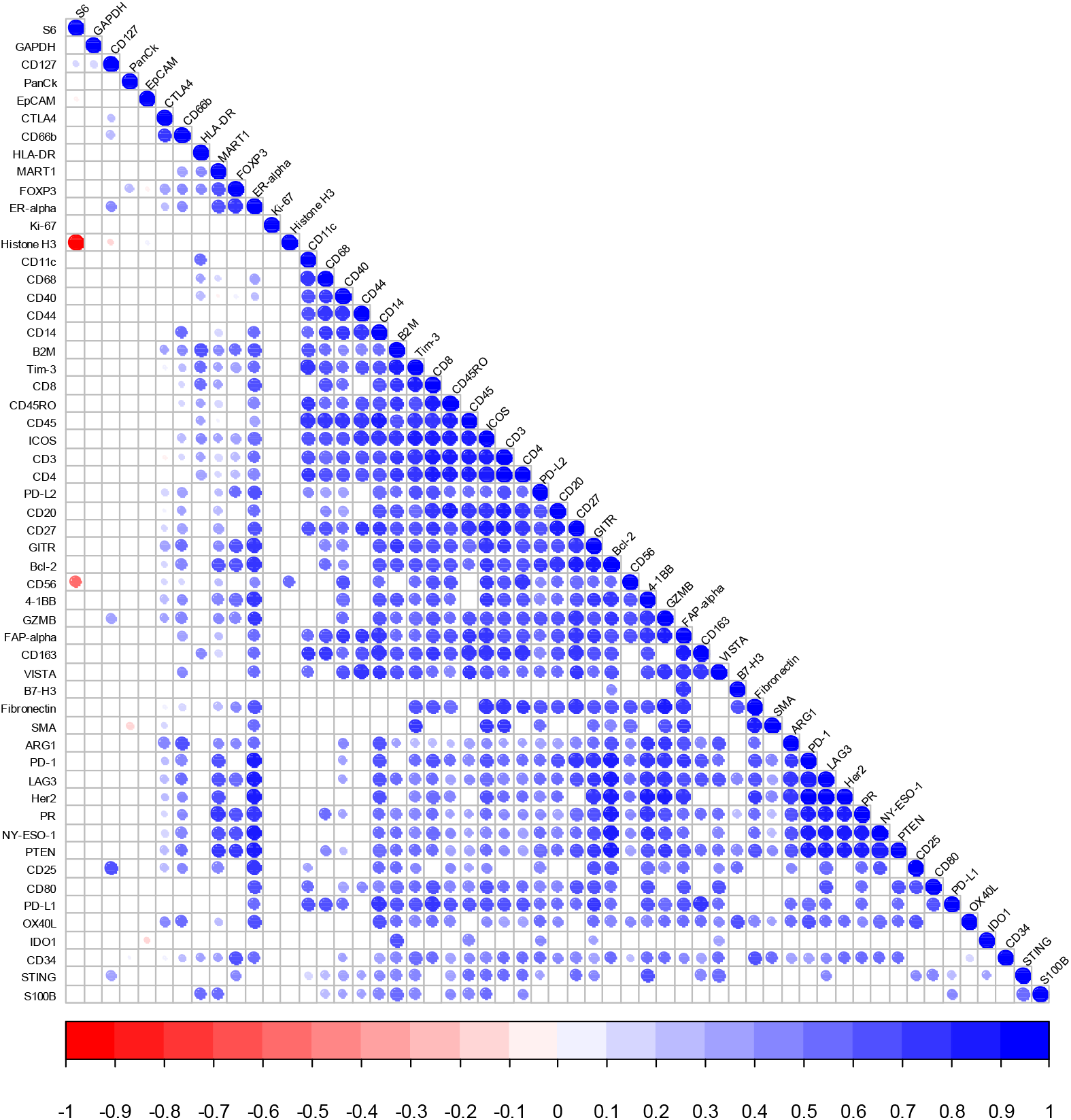
Pearson correlation matrix of proteins within tumour ROIs. Log2 protein expression was used to evaluate Pearson correlations between proteins. Only correlations with p value <0.001 are shown. Blue= positive correlation, Red= negative correlation.

